# "Obesity and metabolic abnormalities: differential associations with subclinical atherosclerosis”

**DOI:** 10.1101/2025.03.29.25324870

**Authors:** Sergio González, Máximo Schiavone, Federico Piñero, Renzo Melchiori, Noelia Brenzoni, Guido García, Pamela Alarcón, Fabián Ferroni, Sergio Barata, Carlos Castellaro

**Author notes:** **Corresponding author, contact information** Sergio González, MD. Department of Cardiology, Hospital Universitario Austral, Argentina Presidente Perón 1500, B1635, Derqui, Argentina.

## Abstract

**Background & Aims:** Obesity is associated with an increased risk of atherosclerosis, though recent evidence shows conflicting results. This study aimed to evaluate whether obesity or its associated metabolic abnormalities play a more significant role in atherosclerosis development in a primary care population.

**Methods:** A cross-sectional study using data from the Cardiometabolic Risk Factor Registry (CARFARE) at Hospital Universitario Austral included adults undergoing their first healthcare visit for primary cardiovascular prevention. Participants were classified into four groups: metabolically healthy non-obese (MHNO), metabolically healthy obese (MHO), metabolically unhealthy non-obese (MUNO), and metabolically unhealthy obese (MUO), according to the BioShare-EU criteria and body mass index. Metabolic abnormalities (MAs) were defined by the same criteria. Atherosclerosis prevalence was analyzed using univariate analysis and multivariable logistic regression models.

**Results:** Among 6,735 participants, 23.3% were MHNO, 3.13% MHO, 45.6% MUNO, and 25.9% MUO. MHO subjects were 10.1% of the obese population. In univariate analysis, atherosclerosis prevalence was higher in obese than non-obese individuals (57.1% vs. 52.0%, p=0.001), but lower in MHNO and MHO compared to MUNO and MUO groups (33.1% and 34.4% vs. 60.4% and 59.5%, p<0.0001). In multivariate regressions, these latter groups presented an increased adjusted odds ratio (aOR) of atherosclerosis compared to MHNO, while atherosclerosis prevalence was no different between the MHO and MHNO groups [aOR: 0.79 (95% CI 0.55-1.11)]. Moreover, in a second logistic regression model, MAs were independently associated with atherosclerosis [aOR: 1.84 (95% CI: 1.60-2.11)], while obesity was not [aOR: 0.91 (95% CI: 0.79-1.04)].

**Conclusion:** In this primary care population, the MHO phenotype was not associated with increased atherosclerosis. MAs, rather than obesity alone, were independently associated with atherosclerosis. These findings highlight the need for further longitudinal studies to clarify the interactions between obesity and metabolic health in atherosclerosis development.

## INTRODUCTION

The World Health Organization (WHO) defines obesity (OBE) as abnormal or excessive fat accumulation that presents health risks [1]. Epidemiological studies have demonstrated that OBE is associated with pathologies such as stroke [2] and ischemic heart disease. Notably, in a collaborative analysis of 57 prospective studies involving 900,000 adults, body mass index (BMI) emerged as a robust predictor of overall mortality. This is primarily attributed to vascular disease [3], which is largely assumed to be atherosclerotic.

In the last 15 years, some studies have suggested a paradoxical relationship between obesity and atherosclerosis (ATS), showing a lower prevalence of aortic and coronary plaque in OBE subjects [4,5]. Moreover, a metabolic healthy obese (MHO) condition has been recently defined [6,7,8,9], which could serve as a category to distinguish whether OBE alone or its accompanying metabolic alterations (MAs) might be associated with the development of subclinical ATS [10]. A better understanding of the mechanism responsible for OBE-associated ATS is essential for refining cardiovascular (CV) prevention strategies.

Little is known about the relationship between OBE and subclinical carotid-ileofemoral ATS in primary CV prevention. Vascular ultrasound to assess carotid-femoral subclinical ATS is a valid approach for stratifying the risk of atherosclerotic CV disease [11]. Using vascular ultrasound to assess subclinical ATS, we were able to compare the differential impact of OBE and associated MA on subclinical ATS development. We conducted a cross-sectional study to determine the prevalence of subclinical ATS in OBE compared to non-obese metabolically healthy (MH) or metabolic unhealthy (MU) individuals. We also investigated whether OBE alone or accompanying MAs is associated with the development of subclinical ATS in subjects accessing CV primary prevention.

## MATERIALS AND METHODS

We conducted a cross-sectional analysis with data from a nested prospective cohort from the Cardiometabolic Risk Factor Registry (CARFARE; Clinical Trials NCT04040777) an integral component of a cardiovascular prevention program, which was conducted at the Cardiometabolic Centre from the Hospital Universitario Austral, Argentina. Our center serves a suburban population around the city of Pilar, located 50 km northwest of the city of Buenos Aires. This population primarily consists of middle-aged individuals of predominantly European descent and of a medium-high economic income. Briefly, this program encompasses sequential evaluations involving a collection of biological sex data, CV risk factors, anthropometric measurements, laboratory determinations of metabolic variables, electrocardiograms, blood pressure (BP) and heart rate measurements, and screening of subclinical ATS in carotid and iliofemoral territories via high-resolution ultrasonography. The study followed STROBE guidelines for reporting observational studies [12]. It complied with international ethical statements and standards of Good Clinical Practice, according to the Declaration of Helsinki, and was approved by the institutional Ethics Committee (CIE 19-044; CARFARE registry).

### Patient Selection

This study entailed analysis of the CARFARE, comprising adult patients who attended our Cardiometabolic Centre between January 2015 and November 2023. We included consecutive adult patients who met the following eligibility criteria: 1) subjects who attended for the first time to undergo a CV evaluation; 2) between 18 and 80 years of age; 2) those who performed a complete evaluation including laboratory analysis and carotid-iliofemoral ATS scanning; 3) in asymptomatic status (without chest pain or any symptom suggestive of cardiovascular disease). Patients were excluded if they had a prior history of coronary artery disease (CAD), including Stable Chronic Angina, Unstable Angina, or Myocardial Infarction; prior Stroke; Chronic Kidney Disease (CKD) defined as glomerular filtration rate < 60 mL/min/m^2^; or Heart Failure (HF).

### Clinical Data, Laboratory Determinations, and CV Risk Factor Assessment

All individuals enrolled in the cardiovascular prevention program underwent physical examination that measured their weight and height to calculate their BMI (weight [kg]/height [m^2^]). Following a 12-hour fast, participants had laboratory examinations, which included plasma glucose (GLU) (UV-Hexokinase enzymatic method), total cholesterol (TC), high-density lipoprotein cholesterol (HDL-C), and triglycerides (TG) levels (enzymatic method). Low density lipoprotein cholesterol (LDL-C) was calculated by the Friedewald method: LDL-C = TC - HDL-C - (triglycerides/5).

The CV risk factors, which included past medical history, physical examination, and laboratory data, were then evaluated. Systemic hypertension [13]; diabetes (DBT) [14]; current smoking [15]; OBE [1]; sedentary habit [16]; dyslipidemia (DLP) [17]; and familial history of CV disease (CVD-FH) [18] were defined following international consensus definitions. Simultaneously, BP was obtained following a 3-minute rest period (Omron HEM 7120). All data were captured prospectively in a database for subsequent analysis.

### Subclinical Atherosclerosis Evaluation

Subclinical ATS screening was conducted in all subjects via B-mode ultrasound and color Doppler evaluation of carotid and femoral bifurcations using a high-resolution vascular ultrasound system (Phillips HD7 XE, Koninklijke Philips N.V) equipped with a 10 MHz linear array probe. Following the Mannheim Consensus, an atherosclerotic plaque was defined as a focal protrusion with a thickness >0.5 mm in the arterial lumen, occupying >50% of the thickness of the surrounding intima-media, or a diffuse thickness >1.5 mm between the media-adventitia and intima-lumen interfaces [19]. ATS plaques were assessed at ten vascular sites: internal, external, and common carotid arteries; iliac external/femoral common arteries; external iliac and superficial femoral arteries. ATS was defined as the presence of ≥1 ATS plaque in the sum of the ten vascular territories analyzed and was independently assessed from exposed and unexposed clinical definitions of subgroup of patients

### Exposure Variables of Interest: Definition of Exposed and Unexposed Study Groups

Patients were classified as OBE or non-obese (NOBE) based on a 30 kg/m² BMI [1]. For the analysis of this cohort, four metabolic groups were defined to address the clinical research question, using the recently published MHO harmonization criteria from the BioShare-EU project as a classification tool [20]. According to this guide, patients in the MHO group must exhibit a BMI > 30 kg/m² in the absence of the following exposure variables: systolic blood pressure (SBP) > 130 mm Hg and/or diastolic blood pressure (DBP) > 85 mm Hg or use of antihypertensives, GLU> 110 mg/dL or use of antidiabetic medications, HDL-C < 40 mg/dL in men or < 50 mg/dL in women or treatment, TG > 150 mg/dL or medication for elevated TG, and a history of CV disease diagnosis. The rest of the OBE subjects were classified as the metabolic unhealthy obese (MUO) group. Similarly, the absence of any MA established by the BioShare-EU project criteria was used in non-obese subjects to define them as the metabolic healthy non-obese (MHNO) group. Non-obese subjects who presented at least one of the MAs pointed out in the BioShare-EU project criteria were classified as part of the metabolic unhealthy non-obese (MUNO) group. In summary, two of the four final groups had MAs (MUNO and MUO), and the remaining two were composed of subjects free of MAs (MHNO and MHO).

### Other Co-Variables

As previously explained, a thorough interrogation was performed to determine the presence of CV risk factors. Since the four study groups were defined by the MHO harmonization criteria of the BioShare-EU project -which involves information on blood pressure, diabetes status, and different MAs-variables such as GLU, HDL-C,TG, LDL-C, hypertension, DBT, and DLP were not taken into account as adjustment variables in the multivariate model to avoid collinearity.

### Statistical Analysis

The analysis was conducted using Stata^®^ 17 OBE. A *p*-value < 0.05 was considered statistically significant. Continuous data are shown using mean (±standard deviation -SD) or median (interquartile range 25-75% - IQR) and were compared using T-test or Mann-Whitney test depending on their distribution, respectively. Categorical data are shown using frequencies or proportions and compared with the Chi-square test. The analysis of differences between more than two groups of continuous variables was performed using ANOVA for those with a normal distribution and the Kruskal-Wallis test for those with a non-normal distribution.

To address the probability of atherosclerosis, multivariable logistic regression models were conducted to estimate odds ratios (OR) and 95% confidence intervals (95% CI). In the multivariable models, we incorporated variables with *p*-values <0.2 in the bivariate analysis (Wald test) on a step-by-step process, following clinical knowledge and biological plausibility (age, sex and CV risk factors) to address confounding effects (change in crude OR >20%). The performances of the final models were evaluated through calibration (Hosmer-Lemeshow test) and discrimination power through the area under the receiving operator curve (AUROC). In the first multivariable logistic regression, we used the four metabolic groups (MHNO, MHO, MUNO, and MUO) as a dummy exposure variable to investigate the independent contribution of each in the development of ATS adjusted for potential confounders. The second logistic regression model used OBE and MAs as exposure variables to investigate their independent effects on ATS development.

## RESULTS

Of the 6,916 subjects having their first CV evaluation, 181 patients with pre-existing CAD, stroke, CKD, or HF were excluded. The remaining 6,735 subjects that entered the study were healthy subjects undergoing primary CV prevention. The participants had a median age of 52 years and were predominantly male (Figure 1). Among them, 4640 (68.9%) were classified as non-obese, and 2095 (31.1%) were classified as OBE. The study population’s detailed demographic and clinical characteristics are presented in Table 1. When compared to non-obese subjects, OBE subjects had significantly worse metabolic health markers, higher BP levels, and a higher prevalence of diabetes (Supplementary Table 1).

**Figure 1:**
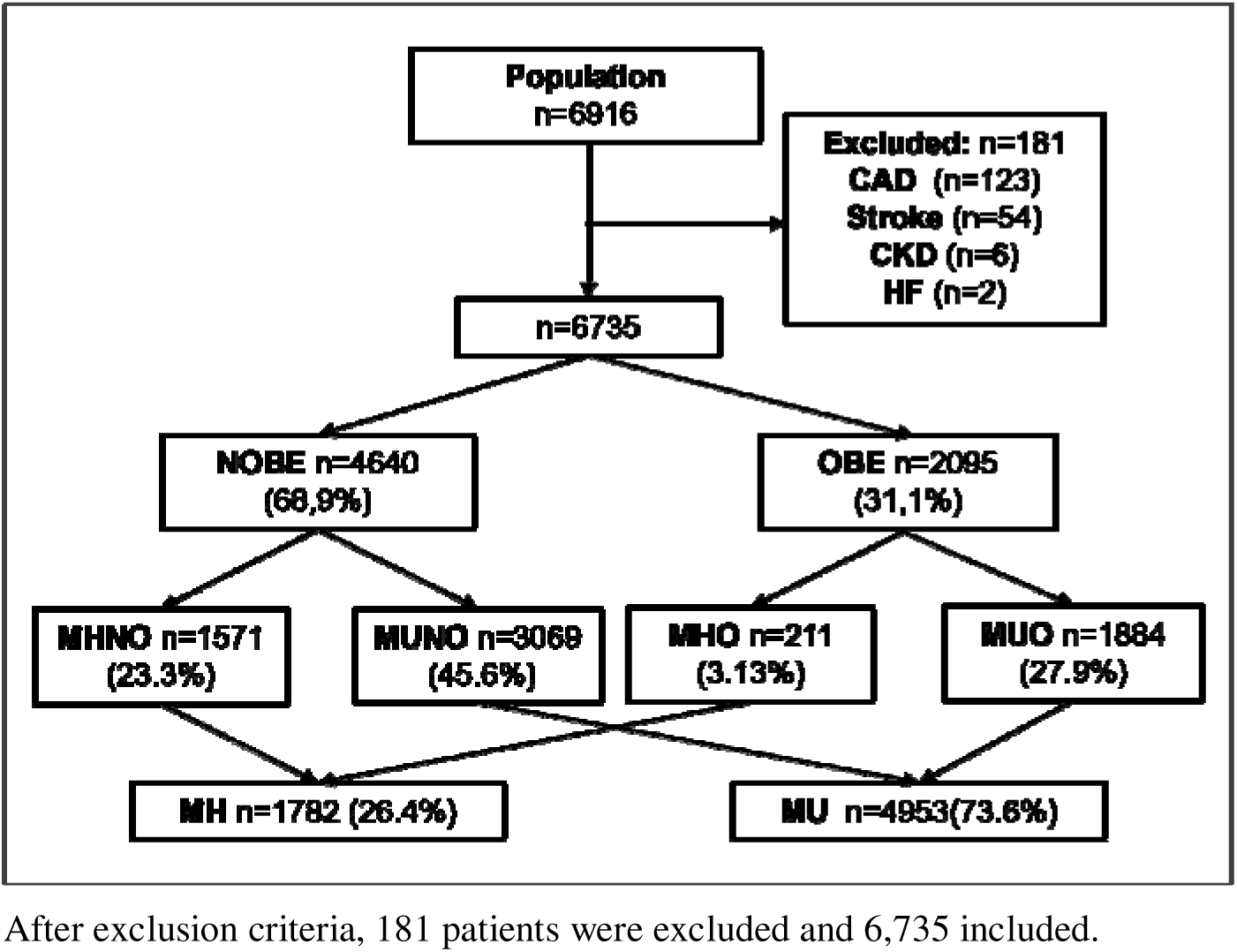
Study population.

**Table 1:** Baseline demographic and laboratory characteristics of included subjects Note: Age, anthropometrics, metabolic variables and smoking habit in the entire population

| VARIABLE | n=6735 |
| --- | --- |
| Age, years (median/IQR) | 53.0 (45-61) |
| Male sex, % | 57.2 |
| BMI, kg/m2 (median/IQR) | 28.4 (25.2-32.4) |
| NOE, % | 68.9 |
| OBE, % | 31.1 |
| SBP, mm Hg (median/IQR) | 131 (122-140) |
| DBP, mm Hg (median/IQR) | 85 (78-91) |
| GLU, mg/dL (median/IQR) | 98.0 (92-106) |
| HDL-C, mg/dL (median/IQR) | 48.0 (41-57) |
| TG, mg/dL (median/IQR) | 122 (88-171) |
| DBT, % | 6.19 |
| Smoking Habit, % | 13.5 |

### Characteristics of MHNO, MHO, MUNO, and MUO groups

Following the classification criteria established by the BioShare-EU project, 23.3% of the subjects were categorized as MHNO, 3.13% as MHO, 45.6% as MUNO, and 25.9% as MUO. The prevalence of MHO among the total number of OBE subjects was 10.1%.Unlike the other groups, the MHNO group was majority female. When we compared the four groups, the metabolic profile progressively worsened from the reference group -MHNO-in the following order: MHO, MUNO, and MUO. They showed sustained increases in BP, GLU, TGs, and a reduction in HDL-C levels. The differences were significant among each group (Table 2). As expected, the MH subjects showed a better metabolic profile than the MU counterparts, which consisted of subjects of predominantly male sex and older age (Supplementary Table 2).

**Table 2:** Comparative analysis between subjects presenting within MHNO, MHO, MUNO and MUO groups.

| VARIABLE | MHNO<br>(n=1571) | MHO<br>(n=211) | MUNO<br>(n=3069) | MUO<br>(n=1884) | P |
| --- | --- | --- | --- | --- | --- |
| Age, years (median/IQR) | 48.5 (42-55) | 48 (42-54) | 54 (46-62) | 52 (44-60) | <0.0001<sup>a</sup> |
| Male sex, % | 36.3 | 64.6 | 56.6 | 74.8 | <0.0001<sup>b</sup> |
| BMI, kg/m<sup>2</sup> (median/IQR) | 23.6 (20.9-25.8) | 32.4 (30.8-34.6) | 25.9 (23.5-27.8) | 33.9 (31.6-37.8) | <0.0001<sup>a</sup> |
| SBP, mm Hg (median/IQR) | 114 (107-120) | 118 (112-124) | 130 (120-139) | 132 (124-142) | <0.0001<sup>a</sup> |
| DBP, mm Hg (median/IQR) | 73 (67-78) | 76 (71-80) | 84 (77-90) | 86 (80-93) | <0.0001<sup>a</sup> |
| GLU, mg/dL (median/IQR) | 92.0 (87-97) | 95.0 (90-101) | 97.0 (91-104) | 100 (94-110) | <0.0001<sup>a</sup> |
| HDL-C, mg/dL (median/IQR) | 60.0 (53-69) | 54.0 (47-61) | 50.0 (43-60) | 45.0 (38-51) | <0.0001<sup>a</sup> |
| TG, mg/dL (median/IQR) | 80.0 (63-101) | 94.0 (72-115) | 112 (82-160) | 138 (102-185) | <0.0001<sup>a</sup> |
| DBT, % | 0.00 | 0.00 | 5.90 | 11.5 | <0.0001<sup>b</sup> |
| Smoking Habit, % | 14.1 | 9.40 | 15.1 | 15.2 | 0.0238<sup>b</sup> |

a. Kruskal Wallis test.
b. Chi-Square test
4 Age, anthropometrics, metabolic variables and smoking habit in the four metabolic groups: Metabolic 5 Healthy Non-obese, Metabolic Healthy Obese, Metabolic Unhealthy Non-obese and Metabolic Unhealthy 6 Obese

### Subclinical Atherosclerosis Prevalence Stratified by Obesity Status and Metabolic Groups

According to the application of the Manheim Criteria, the overall prevalence of carotid/iliofemoral atherosclerosis in our population was 52.7%. Regarding vascular outcomes, in univariate analysis, the prevalence of ATS was higher in OBE subjects than the non-obese ones (57.1% vs. 52.0%, p=0.001, figure 2). However, when the four metabolic groups were considered, ATS was fundamentally associated with metabolic abnormalities. In fact, ATS prevalence in MUNO and MUO was almost doubled than MHNO and MHO groups (Figure 3). Similarly, when only comparing metabolic groups, ATS presence was significantly higher in MU respect MH individuals (59.7% vs. 33.3 %, *p* < 0.0001, Supplementary Figure 1).

**Figure 2:**
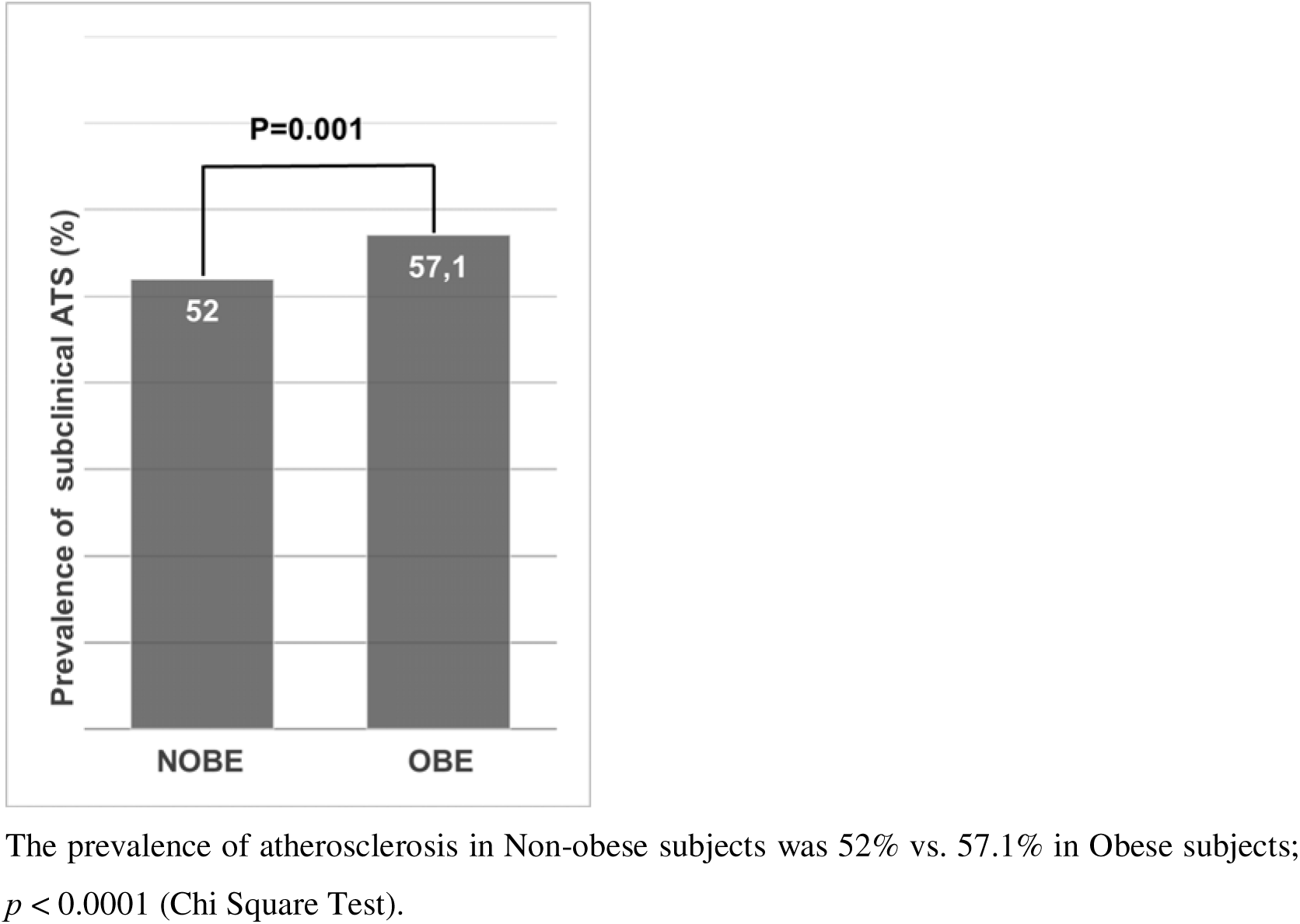
Prevalence of subclinical ATS between NOBE and OBE subjects.

**Figure 3:**
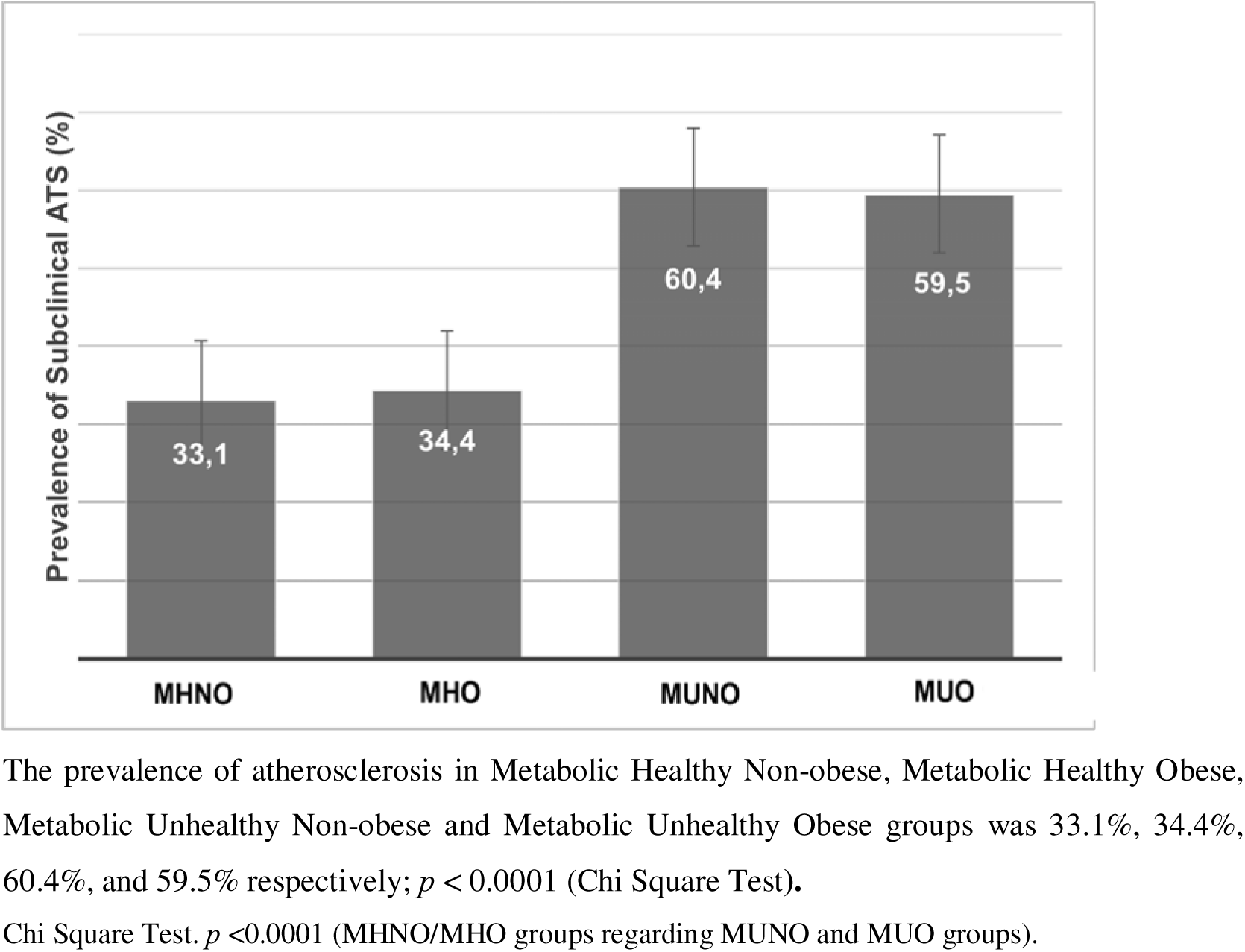
Prevalence of subclinical ATS in MHNO, MHO, MUNO and MUO groups.

### Independent Variables Associated with Subclinical Atherosclerosis

In the first adjusted logistic regression, the odds of having ATS in the MHO group was not statistically different from the reference, the MHNO group [aOR 0.79 (95% CI 0.55-1.11)]. The two MU groups, instead, had higher associations with subclinical ATS than their respective MH group. When adjusted for age, sex, sedentary habit, and smoking, the MUNO and MUO groups were associated with higher odds of having subclinical ATS than the MHNO group [aOR 1.80 (1.54-2.09)] for MUNO, and [aOR 1.68 (1.41-2.00)] for MUO groups, respectively (Table 3). The model performed well, without significant differences in expected and observed ATS proportions (Hosmer & Lemeshow test *p* = 0.32). It had robust power of discrimination, with a c-statistic of 0.83 (95% CI 0.82 - 0.84, Supplementary Figure 2).

**Table 3:** Independent variables associated to ATS presence in logistic regression model 1.

| Variable | Unadjusted<br>OR (95% CI) | P values | Adjusted<br>OR (95% CI) | P values |
| --- | --- | --- | --- | --- |
| **Age** | 1.12 (1.12-1.13) | <0.0001 | 1.13 (1.12-1.14) | <0.0001 |
| **Male sex** | 2.50 (2.30-2.70) | <0.0001 | 3.32 (2.92-3.78) | <0.0001 |
| **Sedentary Habit** | 1.41 (1.27-1.56) | <0.0001 | 1.13 (0.99-1.29) | 0.0614 |
| **Smoking** | 1.57 (1.40-1.80) | <0.0001 | 2.30 (1.93-2.73) | <0.0001 |
| **CVD-FH** | 0.98 (0.86-1.12) | 0.7560 | 1.08 (0.92-1.27) | 0.3282 |
| **Metabolic Groups** |  |  |  |  |
| **MHNO** | Ref. | Ref. | Ref. | Ref. |
| **MHO** | 0.46 (0.34-0.61) | <0.0001 | 0.79 (0.55-1.11) | 0.1819 |
| **MUNO** | 1.76 (1.60-1.94) | <0.0001 | 1.80 (1.54-2.09) | <0.0001 |
| **MUO** | 1.40 (1.26-1.56) | <0.0001 | 1.68 (1.41-2.00) | <0.0001 |

Model 1: $P < .0001$ , $R^2 = 0.42$ (Nagelkerke), Hosmer & Lemeshow test $P = 0.32$ ; AUC (ROC) = 0.83 (95% CI 0.82 - 0.84) In this logistic regression model, atherosclerosis was defined as the dependent variable. We explored the Metabolic Healthy Obese, Metabolic Unhealthy Non-obese, and Metabolic Unhealthy Obese groups (referenced to the Metabolic Healthy Non-obese group) as independent predictors of atherosclerosis adjusted by age, sex, sedentarism, smoking habit, and family history of cardiovascular disease. Model 1: $p$ $< .0001$ , $R^2 = 0.42$ (Nagelkerke), Hosmer & Lemeshow test $p = 0.32$ ; AUC (ROC) = 0.83 (95% CI 0.82 - 0.84)

In the second multiple regression, MAs showed an independent relationship with the presence of ATS [aOR 1.84 (1.60-2.11)] besides age, male sex, and smoking (Supplementary Table 3). Obesity, however, showed no association with subclinical ATS [aOR 0.91 (0.79-1.04)]. This model again performed well, with an adequate calibration (Hosmer & Lemeshow test P= 0.36) and discrimination, with a c-statistic of 0.83 (95% CI 0.82 - 0.84, Supplementary Figure 3).

## DISCUSSION

The present study’s findings suggest that while OBE is associated with a higher prevalence of subclinical ATS compared to non-obese individuals, this relationship is fundamentally influenced by MAs and the presence of accompanying CV risk factors. This conclusion is supported by our observation that the proportion of individuals with MHO who have ATS is comparable to that of their MHNO counterparts. Furthermore, multivariate models revealed no significant differences in the relationship between ATS and MHO or MHNO phenotypes, whereas individuals with MUO and MUNO groups showed stronger associations with ATS compared to the reference MHNO group. Importantly, MAs, rather than obesity itself, emerged as key predictors of ATS in the study population. This suggests that MAs -and likely their triggers, such as insulin resistance (IR)-could be central to the development of ATS in obese individuals.

The concept that not all individuals with obesity share the same risk for developing type 2 diabetes and cardiovascular disease was first introduced by Jean Vague in 1956. Vague attributed these differences to variations in body fat distribution [21]. Later, McLoughlin et al. demonstrated significant heterogeneity in the burden of CV risk factors among moderately obese individuals, particularly linked to variations in IR levels [22]. In their study, patients in the highest IR tertile exhibited higher systolic and diastolic blood pressure, elevated fasting glucose and 2-hour oral glucose load concentrations, higher plasma triglycerides, and lower high-density lipoprotein cholesterol, as well as more prevalent impaired glucose tolerance. Similar findings were reported by Stefan et al., who linked higher liver fat content and abdominal (particularly visceral) adiposity to the MUO phenotype. Conversely, preserved insulin secretion, greater insulin sensitivity, and higher cardiorespiratory fitness were associated with the MHO phenotype [23].

While the number of studies on MHO has grown significantly in recent years [24,25,26], the field lacks consensus on several key issues, the most prominent of which is the definition of MHO. Some studies define MHO as the absence of metabolic syndrome [27], while others offer alternative definitions [7,8,9], leading to substantial variability in reported MHO prevalence across different studies. In the current study, applying the metabolic criteria from the BioShare-EU project, we identified a significant subgroup of approximately 10% of obese individuals (similar to the 12% prevalence observed in the European population) who did not exhibit accelerated atherosclerosis, in contrast to the MUNO and MUO groups.

Recent research has suggested that an early state of vascular insulin resistance may impair muscle and peripheral fat perfusion, leading to whole-body insulin resistance [28]. This alteration in vascular homeostasis could contribute not only to metabolic abnormalities but also to ectopic fat deposition, ultimately promoting the development of ATS. These findings align with the lipid overload and overflow theory proposed by Robert Unger over two decades ago [29]. According to Unger’s theory, far from being purely detrimental, IR might serve as a protective response to reduce lipid-induced tissue damage during short-term overfeeding. By excluding glucose from cells, IR limits glucose-derived lipogenesis. However, with chronic overfeeding, the inability of adipose tissue to expand -partly due to IR and reduced adipose tissue perfusion-leads to the development of ectopic fat deposits in the liver, muscles, and blood vessels. Recent studies have observed this adaptive phenomenon of vascular IR even in non-obese individuals subjected to short-term overfeeding [31], where reduced insulin-induced vasodilation in skeletal muscle resistance arteries shifts tissue perfusion away from muscle and toward adipose tissue. This phenomenon, seemingly transient in nature, may help explain the relative instability of the MHO phenotype. Indeed, MHO is often associated with a shorter duration of obesity [32] and is more common among younger individuals [33], as was also observed in our study.

The present study has both strengths and limitations. A key strength is its inclusion of a large cohort of individuals without clinical cardiovascular disease, all of whom underwent comprehensive metabolic and vascular assessments. This design allowed us to evaluate the differential impact of MAs and CV risk factors on the development of ATS, beyond simple anthropometric measures. However, a notable limitation of our study was the lack of direct measures of body fat distribution, such as waist circumference [34,35] or waist-to-hip ratio [36,37], which have proven to be sensitive markers for metabolic disorders, diabetes [38,39], and CV disease [38,40]. Nevertheless, most definitions of MHO rely on BMI [6,7,8], a readily available metric often used in large cohort studies.

In our analysis of ATS, we did not observe a paradoxical relationship between obesity and subclinical ATS, nor could we establish an independent relationship between the two. These findings suggest that the development of ATS is more closely associated with metabolic abnormalities and accompanying CV risk factors rather than obesity per se. Therefore, early intervention targeting metabolic abnormalities and cardiovascular risk factors could play a critical role in preventing carotid-ileofemoral ATS in obese individuals. Nonetheless, further research is needed to confirm these findings and to explore the underlying mechanisms.

## Data Availability

Data is available upon reasonable request

## Acknowledgements

We would like to thank to the entire staff of the Cardiometabolic Centre (Laura Ayerdi, Adrian Chehda, Cecilia Spaenocchia, Diego Aruffe, Eliana Filosa, Matias Failo, Julieta Bustamante, Susana Apoloni, Victoria Napoli, Ana Bernhardt, Silvina Zappoli) and Jorge Chiabaut Svane for their invaluable collaboration.

## Authors contributions

All the authors approved the final version of the manuscript.

*Concept and study design:* Sergio González.

*Data collection:* Sergio González, Noelia Brenzoni, Guido García, Pamela Alarcón.

*Analysis:* Máximo Schiavone, Sergio González, Renzo Melchiori.

*Writing the article:* Sergio González, Máximo Schiavone, Federico Piñero.

*Critical Revision:* Federico Piñero, Sergio González, Máximo Schiavone, Renzo Melchiori, Sergio Barata, Noelia Brenzoni, Guido García, Pamela Alarcón, Fabián Ferroni, Carlos Castellaro.

## SUPPLEMENTARY MATERIAL

**Supplementary Table 1:** Comparative analysis between NOBE and OBE subjects.

| **VARIABLE** | **NOBE**<br>(n=4640) | **OBE**<br>(n=2095) | ***p*** |
| --- | --- | --- | --- |
| **Age**, years (median/IQR) | 52 (45-60) | 51 (44-59) | 0.0680* |
| **Male sex**, % | 49.3 | 73.8 | < 0.0001<sup>#</sup> |
| **BMI**, kg/m<sup>2</sup> (median/IQR) | 25.2 (22.6-27.4) | 33.7 (31.7-36.9) | < 0.0001* |
| **SBP**, mm Hg (median/IQR) | 123 (113-134) | 130 (122-141) | < 0.0001* |
| **DBP**, mm Hg (median/IQR) | 80 (73-87) | 85 (78-92) | < 0.0001* |
| **GLU**, mg/dL (median/IQR) | 95.0 (89-102) | 100 (93-108) | < 0.0001* |
| **HDL-C**, mg/dL (median/IQR) | 54.0 (46-64) | 45.0 (39-53) | < 0.0001* |
| **TG**, mg/dL (median/IQR) | 98.0 (73-135) | 131 (97-179) | < 0.0001* |
| **DBT**, % | 4.20 | 10.5 | < 0.0001<sup>#</sup> |
| **Smoking Habit**, % | 14.0 | 12.3 | 0.0590<sup>#</sup> |

Note: \*Mann Whitney test.
<sup>#</sup>Chi-Square test
**Note:** Age, anthropometrics, metabolic variables and smoking habit in Non-obese vs. Obese subjects

**Supplementary Table 2:** Comparative analysis between MH and MU groups.

| **VARIABLE** | **MH**<br>(n=1782) | **MU**<br>(n=4953) | ***p*** |
| --- | --- | --- | --- |
| **Age**, years (median/IQR) | 48.0 (42-55) | 53.0 (45-61) | < 0.0001* |
| **Male sex**, % | 39.7 | 63.5 | < 0.0001<sup>#</sup> |
| **BMI**, kg/m<sup>2</sup> (median/IQR) | 24.2 (21.3-27.1) | 28.4 (25.2-32.4) | < 0.0001* |
| **SBP**, mm Hg (median/IQR) | 114 (108-121) | 131 (122-140) | < 0.0001* |
| **DBP**, mm Hg (median/IQR) | 73 (68-78) | 85 (78-91) | < 0.0001* |
| **GLU**, mg/dL (median/IQR) | 92.0 (87-97) | 98.0 (92-106) | < 0.0001* |
| **HDL-C**, mg/dL (median/IQR) | 59.0 (52-69) | 48.0 (41-57) | < 0.0001* |
| **TG**, mg/dL (median/IQR) | 81.0 (64-104) | 122 (88-171) | < 0.0001* |
| **DBT**, % | 0.00 | 9.54 | < 0.0002<sup>#</sup> |
| **Smoking Habit**, % | 13.4 | 13.8 | 0.1920<sup>#</sup> |

Note: \*Mann Whitney test. #Chi-Square test
Note: \*Mann Whitney test.

**Supplementary Table 3:** Independent variables associated to ATS presence in logistic regression model 2.

| Variable | Unadjusted |  | Adjusted |  |
| --- | --- | --- | --- | --- |
|  | OR (95% CI) | *p* | OR (95% CI) | *p* |
| **Age** | 1.12 (1.12-1.13) | < 0.0001 | 1.13 (1.12-1.14) | < 0.0001 |
| **Male sex** | 2.50 (2.30-2.70) | < 0.0001 | 3.32 (2.92-3.78) | < 0.0001 |
| **Sedentarism** | 1.41 (1.27-1.56) | < 0.0001 | 1.13 (1.00-1.29) | 0.0569 |
| **Smoking** | 1.57 (1.40-1.80) | < 0.0001 | 2.30 (1.94-2.73) | < 0.0001 |
| **CVD-FH** | 0.98 (0.86-1.12) | 0.7560 | 1.08 (0.92-1.28) | 0.3209 |
| **OBE** | 1.23 (1.11-1.37) | 0.0001 | 0.91 (0.79-1.04) | 0.1774 |
| **MA** | 2.98 (2.66-3.33) | < 0.0001 | 1.84 (1.60-2.11) | < 0.0001 |

3 Model 2: $P < 0.0001$ , $R^2 = 0.42$ (Nagelkerke), Hosmer & Lemeshow test $P = 0.36$ ; AUC (ROC) = 0.83 (95% CI 0.82- 4 0.84)
**Note:** In this logistic regression model atherosclerosis was defined as the dependent variable, while Obesity and Metabolic Abnormalities were explored as independent predictors of atherosclerosis adjusted by Age, Male Sex, Sedentarism, Smoking Habit and Family History of Cardiovascular Disease.

**Supplementary Figure 1:**
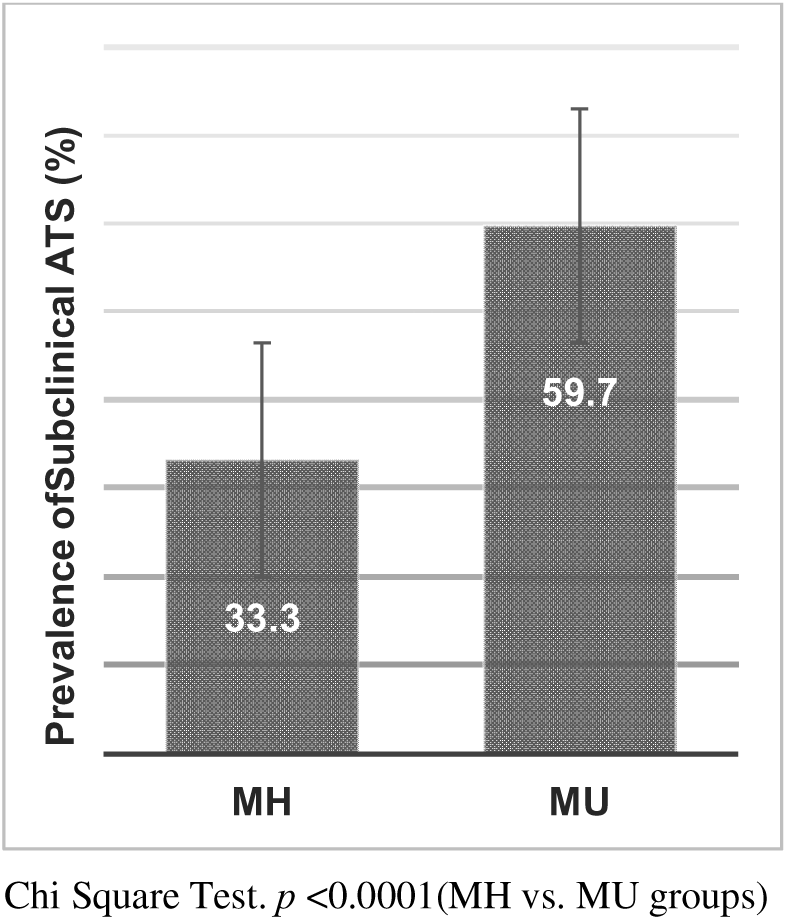
Prevalence of subclinical ATS between MH and MU groups.

**Supplementary Figure 2:**
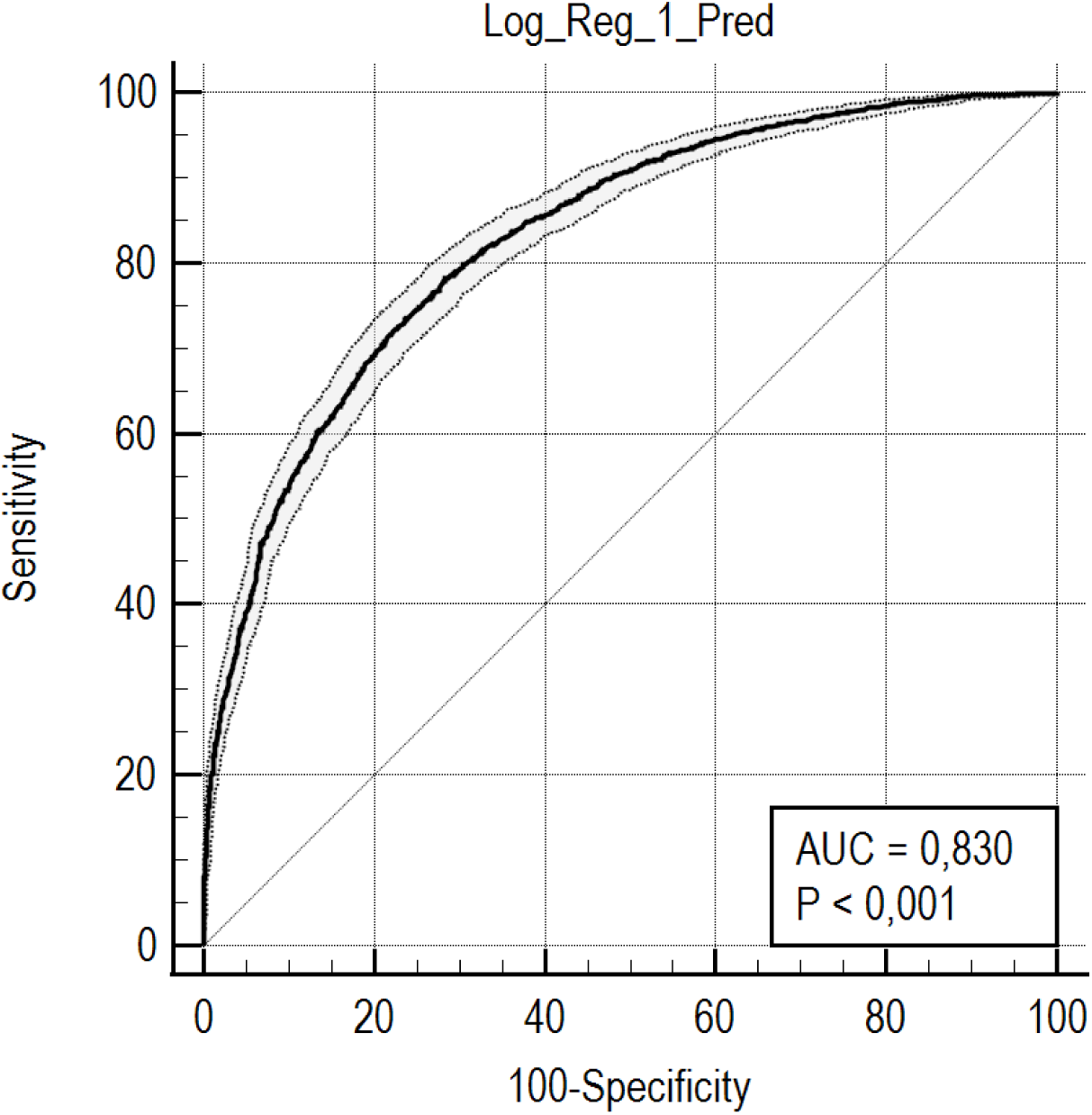
ROC curve of logistic regression model 1.

**Supplementary Figure 3:**
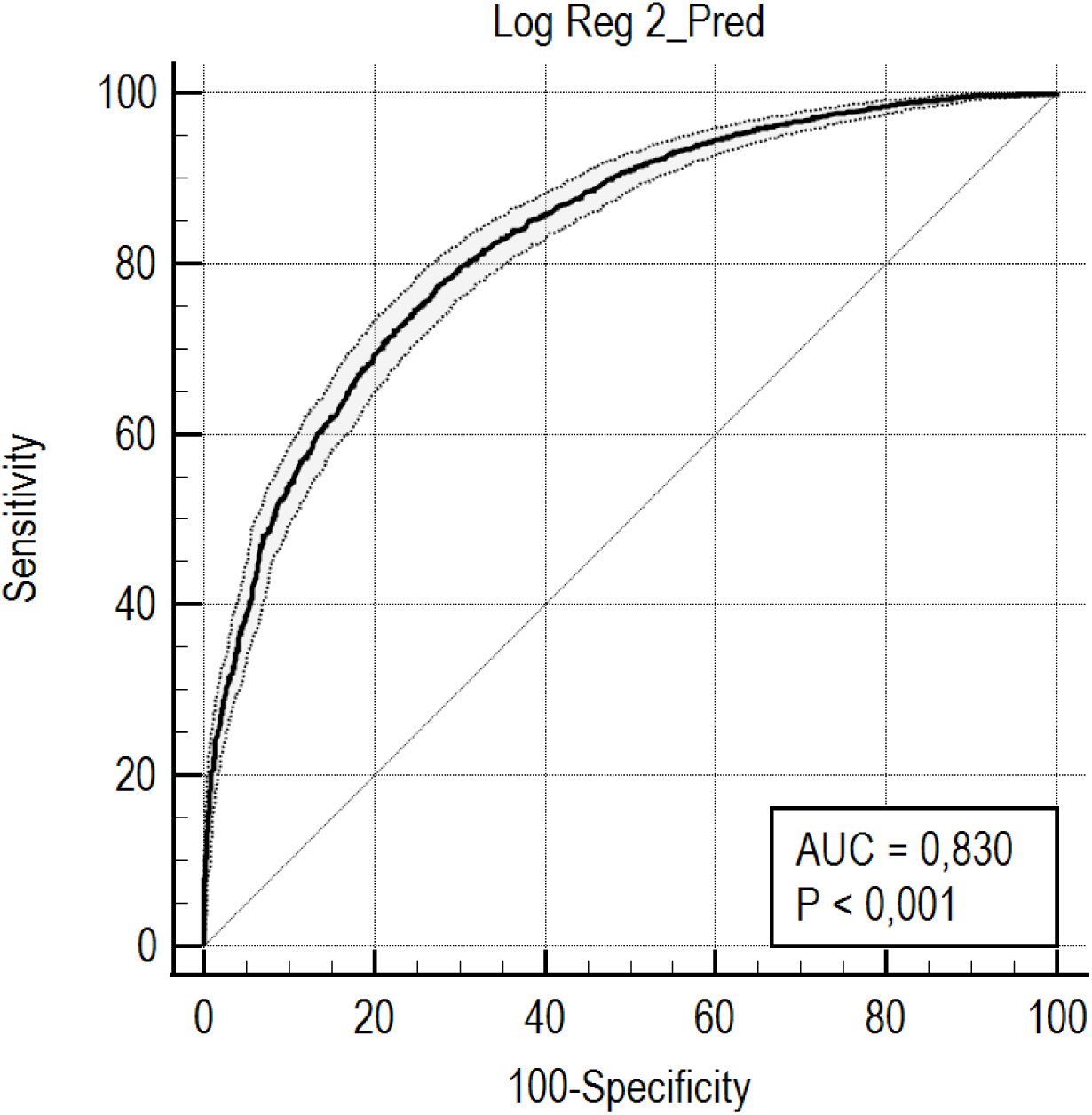
ROC curve of logistic regression model 2.

